# Compromised intestinal barrier underlies gut microbiota dysbiosis of intestinal diseases

**DOI:** 10.1101/19011833

**Authors:** Puzi Jiang, Senying Lai, Sicheng Wu, Xiushan Yin, Jingjing Zhang, Xing-Ming Zhao, Shun-Qing Xu, Wei-Hua Chen

**Affiliations:** Key Laboratory of Molecular Biophysics of the Ministry of Education, Hubei Key Laboratory of Bioinformatics and Molecular-imaging, Department of Bioinformatics and Systems Biology, College of Life Science and Technology, Huazhong University of Science and Technology, 430074 Wuhan, Hubei, China; Huazhong University of Science and Technology Ezhou Industrial Technology Research Institute, 436044 Ezhou, Hubei, China; Institute of Science and Technology for Brain-Inspired Intelligence, Fudan University, Shanghai 200433, China; Applied Biology Laboratory, Shenyang University of Chemical Technology, 110142 Shenyang, China; Key Laboratory of Environment and Health, Ministry of Education & Ministry of Environmental Protection, and State Key Laboratory of Environmental Health, School of Public Health, Tongji Medical College, Huazhong University of Science and Technology, Wuhan, 430030 Hubei, China; Key Laboratory of Computational Neuroscience and Brain-Inspired Intelligence, Ministry of Education, China; College of Life Science, HeNan Normal University, 453007 Xinxiang, Henan, China

## Abstract

**Background:** Despite recent efforts, a single factor underlying the gut microbiota dysbiosis in intestinal diseases is not identified. We hypothesized that compromised intestinal barrier (CIB) could lead to increased host-derived contents including human cells in the gut, change its physio-metabolic properties, and globally alter gut microbiota and their metabolic capacities.

**Results:** Consistently, we found human DNA contents (HDCs), calculated as the percentage of metagenomic sequencing reads mapped to the human genome, were significantly elevated in colorectal cancer (CRC) patients; HDC correlated with microbial- and metabolic-pathway-biomarkers of CRC, and was the most important contributor to patient stratification. We found similar results in Crohn’s disease (CD); additionally, patients treated with diet and drug intervention showed reduced HDC levels over time, and were accompanied by reversing changes of many CD-signature species.

**Conclusions:** Our results suggested that host-derived contents may have greater impact on gut microbiota than previously anticipated, and CIB could be an ideal treatment target that could reverse dysbiosis globally and precisely.

## Background

Colorectal cancer (CRC) is the 3^rd^ most common cancer worldwide and the 2^nd^ leading cause of cancer-related death in the United States [1, 2]; in recent years, the incidence of CRC has been increasing in young adults in major western countries [3, 4]. Similarly, Crohn’s disease (CD) is also increasing worldwide and can be attributed largely to industrial urbanization and Western life-styles [5]. As genetics could only explain limited proportions of the CRC [6, 7] and CD [8] incidences, researchers have recently linked it to environmental factors, life styles and gut microbiota dysbiosis [8-13]. By contrasting gut microbiome profiles of CRC and CD patients to that of the healthy controls, researchers have identified bacterial species that were specifically enriched in CRC [10-12, 14] and CD [13] respectively; many of the CRC-enriched species were recently found to be consistent across populations, according to two meta-analysis studies [15, 16]. In addition, microbial genes involved in various biological pathways were also enriched in the gut microbiota of CRC [10, 15, 16] and CD [13] patients. Both the differential species and pathways can be used as non-invasive markers for patient stratification [10, 11, 13, 15, 16]. These findings greatly improved our understanding on the potential roles of gut microbiota in the pathogenesis and/or development of these intestinal diseases, and implied a global alteration of the local gut environment in the patients. However, a single dominant factor that underlies (e.g. globally correlates with biomarkers of) the gut microbiota dysbiosis associated with these diseases has not been identified.

Compromised intestinal barrier (CIB) has been shown to associate with many intestinal diseases, including inflammatory bowel diseases (IBD) [17] and CRC [18, 19]. CIB could increase the amount of host-derived contents, including epithelial cells and blood to be shed into feces [20], alters the physio-metabolic properties of the gut environment [21] and consequently leads to global alterations in gut microbiota composition (dysbiosis). We thus hypothesized that CIB could be a major factor underlying the gut microbiota dysbiosis in CRC and CD: the severity of which may correlate with the extent of the dysbiosis.

Previous researches have detected increased human DNAs in feces from patients of various diseases using real-time polymerase chain reaction (RT-PCR) technique. For example, patients with inflammatory or pathogenic agents such as *Clostridium difficile* infection were found with higher human DNA content in feces and lower microbial diversity [22, 23]. Since fecal metagenomics are obtained using whole-genome shotgun sequencing and contain unbiased survey on bacterial, viral and host DNA contents, we could directly calculate the human DNA content (HDC) as the percentage of the gut metagenomics sequencing reads mapped to the human genome (see Methods) for each fecal sample and use it as a proxy of CIB.

In this study, we collected eight metagenomic datasets from two most common intestinal diseases. We confirmed our hypothesis that not only HDC was significantly elevated in the patients, but also they were significantly correlated with disease-signature species and metabolic pathways. HDC could improve the performance of patient stratification models, and ranked as the most important contributing factor. More importantly, we found that HDC can be used as a better biomarker for effective treatment, because it signified the global recovery of altered species in Crohn’s disease. Together, our results suggested that host derived contents, as proxied by HDC, had greater impact on gut microbiota than we previously expected; treatment targeting the source of increased host contents, i.e. CIB, can reverse gut dysbiosis in a global and precise manner.

## Results

### Increased human DNA contents (HDCs) in CRC patients

We first focused on CRC. As expected, we found that HDCs were significantly higher in feces of CRC patients than that of the healthy controls in all seven datasets (Fig. 1a, Tables S1 and Tables S2). We identified in total 21 species that were significantly correlated with HDCs in more than two datasets (Spearman Rank Correlation, Fig. 1b; see Methods and Table S3). Among which, eleven overlapped with the CRC microbial signatures identified by the two recent meta-analysis (referred to as CRC-signature species below) [15, 16], including ten CRC-enriched and one CRC-depleted species (Fig. 1b). We also identified sixteen metabolic pathways that were significantly correlated with HDCs (Table S4); among which, five were previously identified metabolic-pathway-biomarkers for CRC (referred to as CRC-signature pathways below). These results validated our hypotheses that CIB, as indicated by HDCs that can be directly quantified from gut metagenomics data, underlies the dysbiosis in CRC patients.

**Fig. 1.**
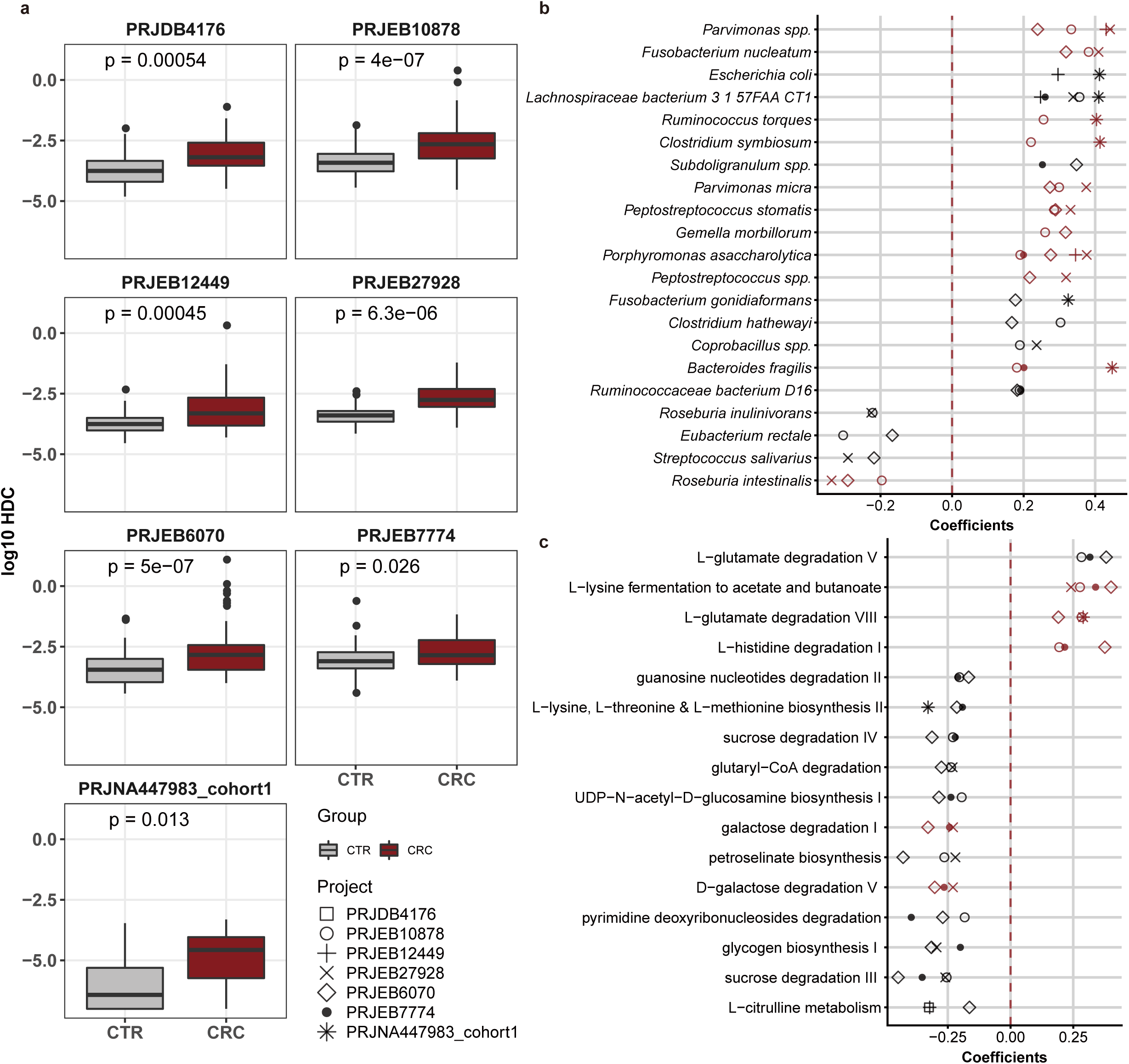
Human DNA contents (HDCs) were significantly elevated in feces of CRC patients, and correlated with microbial- and metabolic-pathway-biomarkers. **a**, HDCs, calculated as the percentage of gut metagenomics sequencing reads mapped to the human genome, were significantly higher in CRC (dark red box) than healthy controls (grey box) in seven recently published datasets (Wilcoxon Rank Sum Test, see Methods). **b**, Species that were significantly correlated with HDCs in two and more CRC datasets (Spearman Rank Correlation, see Methods). Correlations were calculated using both CRC patients and healthy controls. Red: previously identified microbial-biomarkers of CRC [15, 16]. **c**, Metabolic pathways that were significantly correlated with HDCs in three and more CRC datasets. Correlations were calculated using both CRC patients and healthy controls. Red: previously identified pathway-biomarkers of CRC [15, 16].

### Increased HDC underlie altered species and metabolic pathways in CRC and contributed significantly to patient stratification

We next tested if HDC and correlated species and pathways (referred as to HDC-species and HDC-pathways respectively) could contribute to patient stratification in CRC. Similar to Wirbel *et al* [15] and Thomas *et al* [16], we performed a leave-one-dataset-out (LODO) analysis [24] in which Random forest classifiers were trained on the combined datasets of all but one, and tested on the one that was left-out; we did this for each dataset in turn. As shown in Fig. 2a and 2c, for models trained using species and pathways abundances, including HDCs could improve prediction performance. More importantly, HDC was ranked as the most important feature in models trained with HDCs (Fig. 2b and 2d). In addition, in the taxonomic-based models, all of the top ten features were HDC-species (excluding HDC itself if it was used in model training); by contrast, only six were CRC-signature species described by the previous meta-analyses (Fig. 2b, see also ref [15, 16]). Similarly, in the pathway-based models, nine out of the top ten features were HDC-pathways, while six of them were CRC-signature pathways. These results indicated the HDC-correlated features could contribute substantially to patient stratification and diagnosis. It’s worth to note that models trained on HDC-species and differential-species identified using Wilcoxon Rank Sum Test (see Methods) did not differ significantly in their predictive performance (Fig 2a), implying redundant roles of some species in model training; similar results were found in HDC-pathways (Fig 2c).

**Fig. 2.**
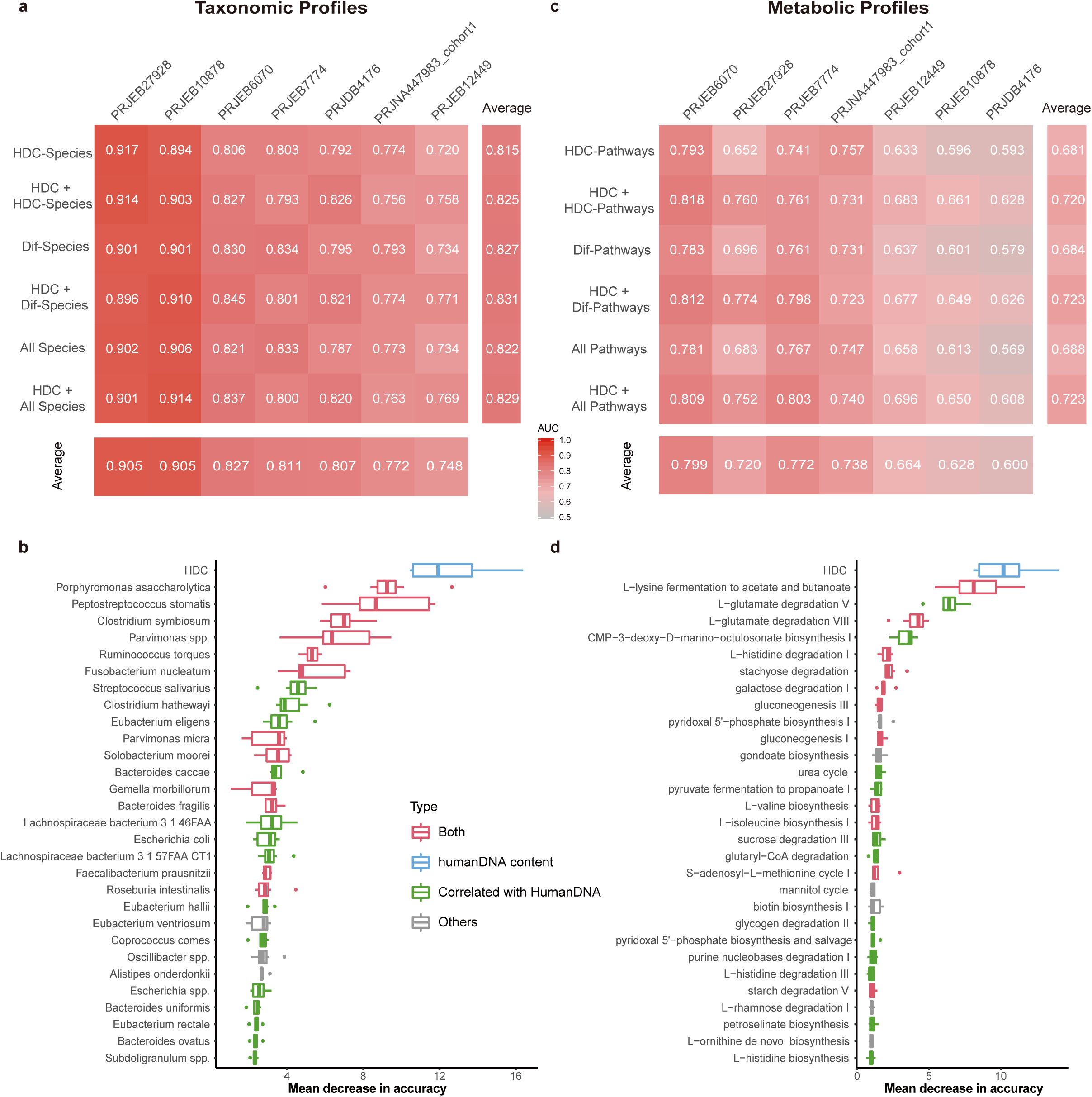
HDC and correlated species and metabolic-pathways contribute significantly to patient stratification in LODO analysis in CRC. **a**, Predictive performances as AUC values obtained using leave-one-dataset-out (LODO) analysis by training the models on the species abundances. The AUC values were averaged from repeated results of 10-fold validation analysis. Dif-species: species whose abundances are significantly different between CRC and controls in at least one dataset (Wilcoxon Rank Sum Test, see Methods); HDC-species: HDC-correlated species; see Methods for details. All-species: models build on all species. **b**, Ranking of feature importance in the HDC + All-species model. The models were trained by using HDC values and relative abundances of all species as input. The importance scores were reported by the LODO models. The features were ranked according to the median importance scores from 100 repeated results of 10-fold validation analysis, see Methods. Blue: HDC, Green: HDC-species; Red: Dif-species that correlated with HDC, Grey: other species. **c**, AUC values obtained using LODO analysis by training the models on the metabolic pathway abundances. Dif-pathways: pathways whose abundances are significantly different between CRC and controls (Wilcoxon Rank Sum Test, see Methods); HDC-pathways: HDC-correlated pathways; see Methods for details. All-pathways: models build on all pathways. **d**, Ranking of feature importance in the HDC + All-pathway model. The models were trained by using HDC values and relative abundances of all pathways as input. The importance scores were reported by the LODO models. Blue: HDC, Green: HDC-correlated pathways; Red: Dif-pathways that correlated with HDC, Grey: other pathways.

### Similar results were found in CD

We then checked if similar results could be found in CD. A previous study reported elevated fecal HDCs in pediatric CD as compared with healthy controls [13]; the authors used quantitative polymerase chain reaction (QPCR) method to quantify HDCs by targeting human beta-tubulin coding-sequences. The authors also calculated HDCs from the metagenomics data and reported that the QPCR results were positively correlated with metagenomics-data-derived HDC values (r = 0.81 Pearson’s correlation, p = 9.3 × 10^−11^; see ref [13]). We re-calculated the HDCs using our methods and found they were highly correlated with theirs (r = 0.977 Pearson’s correlation, p = 4.16 × 10^−109^; Table S5). These results further validated the reliability and accuracy of metagenomics-derived HDCs.

We identified 40 HDC-correlated species, most of which were also differential-species (CD-signature species) that showed significant abundance changes between healthy controls and untreated patients (Control+Baseline group, Fig 3a, Table S6 and Table S7). We also built Random forest classifiers using species abundances for CD and did 20 times repeated 10-fold cross-validation. Similar to CRC, we found that adding HDC to the input data could improve prediction performance (AUC increased from 0.938 to 0.953); also similar to CRC, we found that HDC was ranked as the most important feature, and top ten features were all HDC-correlated (Fig 3b). Interestingly, although overlapped significantly, these species are quite different from those in CRC (Table S8) in terms of their changes and importance in patient stratification (Fig. 3b), likely due to the fact that CD occurred at the small intestine (ileum) and colon, while CRC occurred at more downstream of the intestinal tract. Nonetheless, it appears that elevated HDC is a common feature of intestinal diseases, while different diseases can be distinguished by their different gut dysbiosis profiles.

**Fig. 3.**
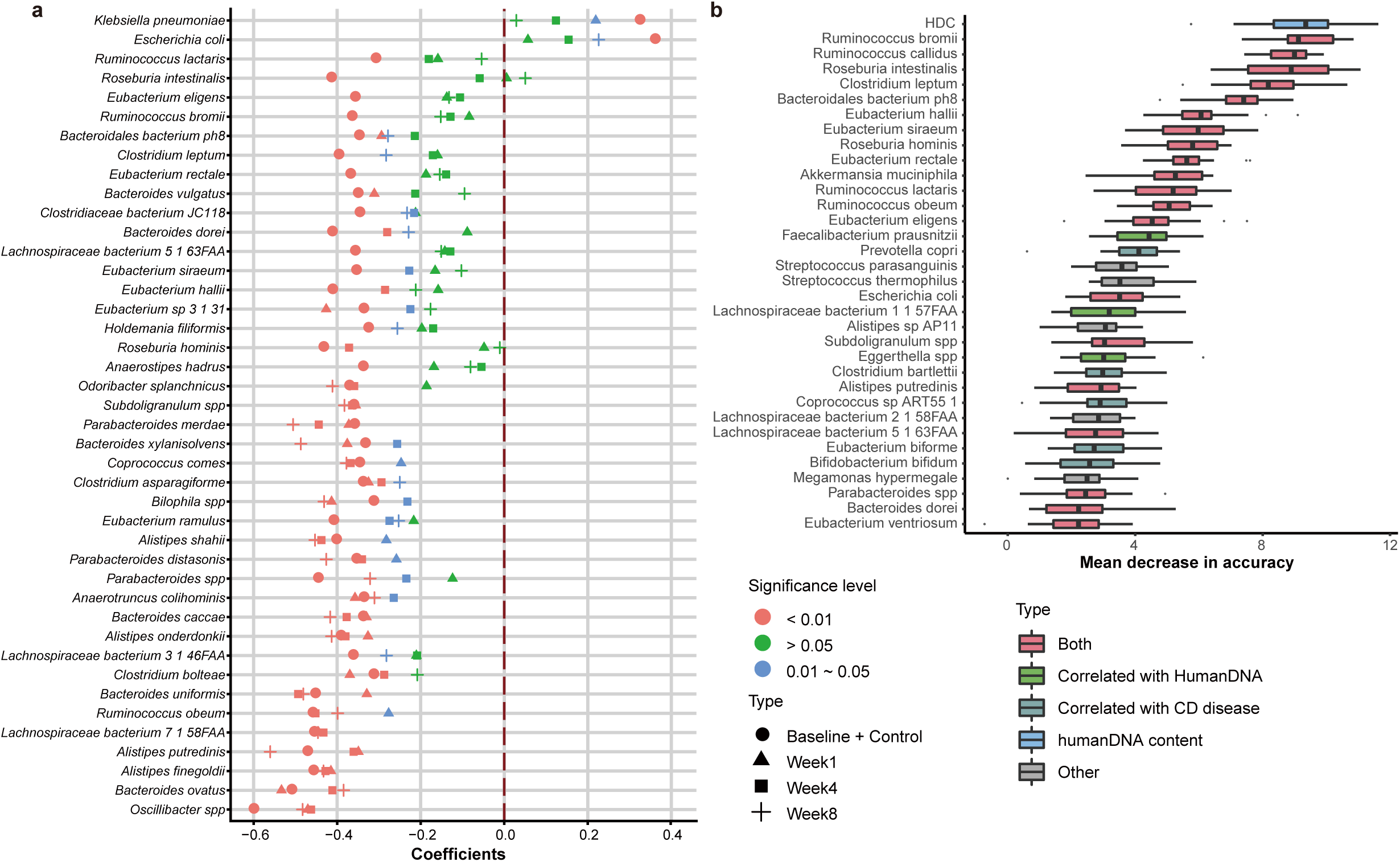
HDC was also elevated in CD, correlated with differential species and contributed significantly to patient stratification. **a**, Species that were correlated with HDCs in the group of healthy controls and untreated patients (Baseline + Control). Also plotted are the correlation coefficients between HDCs and species abundances in patients at three time-points after they were treated (Week1, Week4 and Week8). Correlation coefficients were color-coded according to their significance levels. **b**, Ranking of feature importance in the HDC + All-species model. The models were trained by using HDC values and relative abundances of all species as input; only the data of the healthy controls and untreated patients were used. The importance scores were reported by the Random forest models. The features were ranked according to the median importance scores from 200 repeated results of 10-fold validation analysis, see Methods. Blue: HDC, Green: HDC-species; Red: species whose abundances are correlated with HDC and significantly different between healthy controls and the untreated CD patients, Grey: other species.

### HDC signified clinical treatment outcomes and recovery of disease-altered species

The CD patients were treated with diet intervention and anti-TNF antibodies; the outcomes were evaluated with fecal metagenomics sequencing at week 1, 4 and 8 after the interventions [13]. We found that the HDCs were significantly decreased over time (Fig 4a). Strikingly, we found majority of the HDC-correlated species showed coordinated changes with HDC, i.e. species that were positively (negatively) correlated with HDC in the Control+Baseline group decreased (increased) with the decreasing HDCs (Fig. 4b-f, Figure S1), suggesting that the intervention that reduced fecal HDCs could globally reverse the gut dysbiosis in a species-specific manner. Such a conclusion was further supported by the observation that the correlations between HDC and many of the species were consistent in the Control+Baseline, Week1, Week4 and Week8 groups (Fig 3b). As expected, HDC correlates significantly with fecal calprotectin (FCP; Pearson’s correlation = 0.518, *p* = 1.4× 10^−23^, Figure S2), a clinical indicator of intestinal inflammation. However, the correlations between CD-signature species and FCP were much lower than that of the HDC (Figure S3). These results indicated that HDC is a better biomarker for the disease and effective treatment.

**Fig. 4.**
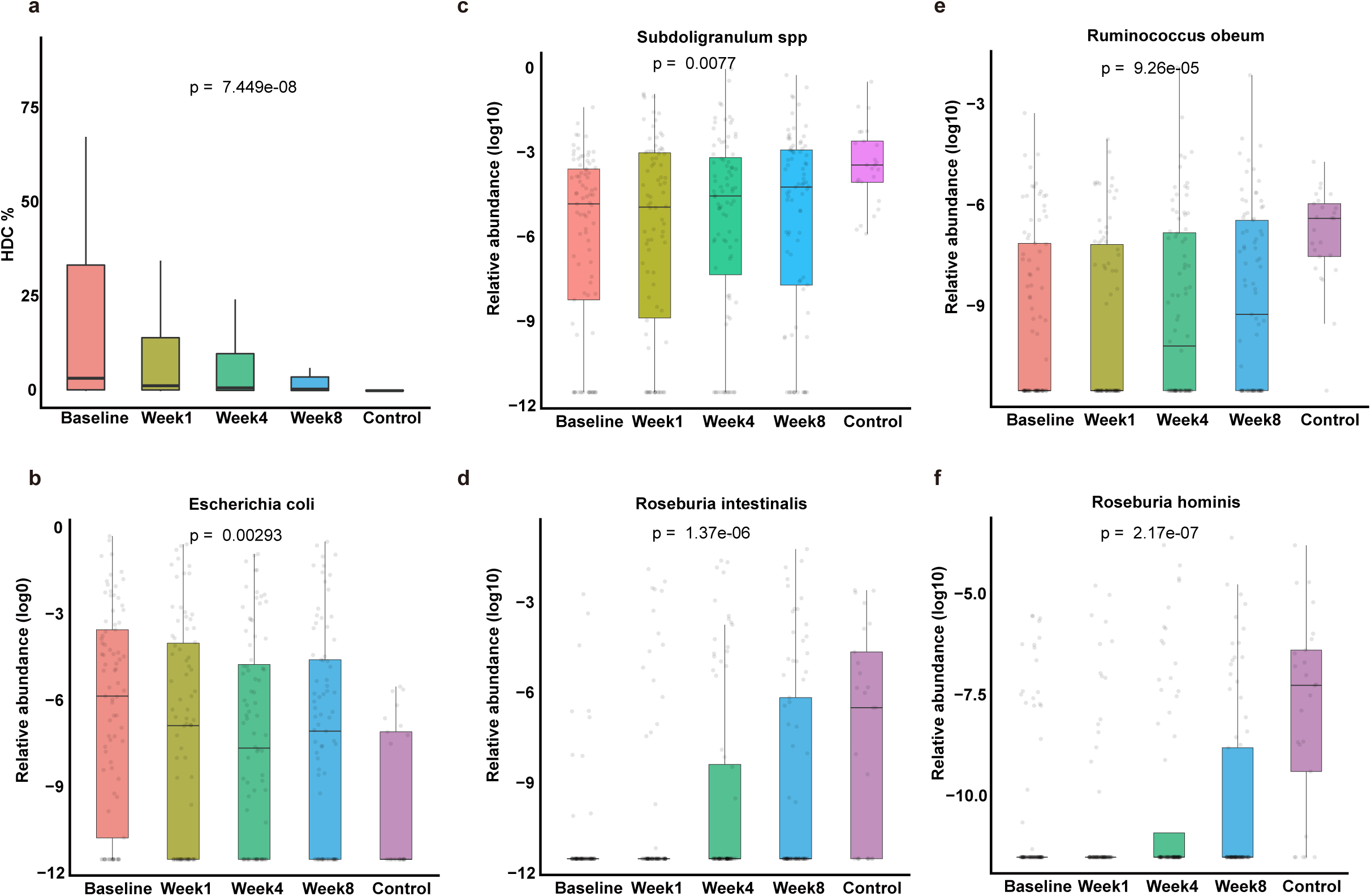
Treatment that reduced fecal HDCs could globally and precisely reverse the changes of HDC-correlated species in CD. **a**, HDCs were significantly reduced along the course of treatment. **b**, The abundance changes of *Escherichia coli* along the course of treatment; p-values were calculated by comparing the abundances between groups (Kruskal-Wallis test). Other exemplary species include: **c**, *Subdoligranulum spp*. **d**. *Roseburia intestinalis*. **e**, *Ruminococcus obeum*. **f**, *Roseburia hominis*.

## Discussion

Together, our hypothesis driven method identified compromised intestinal barrier (CIB), indicated by elevated human DNA contents (HDCs) in feces, as a major factor underlying the gut microbiota dysbiosis in patients with intestinal diseases. CIB can increase the host-derived contents including epithelial cells and/or blood to be shed into feces, alter the local gut environment and lead to gut microbiota dysbiosis. Due to the reciprocal relationship between gut microbiota and the host [25, 26], the latter may lead to more severe CIB conditions. So far researchers have mostly focused on the impact of CIB on the host [27-30], much less on the other way round; our results demonstrated that the increased host-derived contents in the gut due to CIB may have greater impact on gut microbiota that we previously anticipated. Our results suggested that CIB could be an ideal target for treatment and intervention: by targeting the molecular processes that might cause the elevated HDC, and/or reversing the physiological and metabolic changes that CIB brought to the feces, we should be able to first improve the gut local environments and then reverse the gut microbiota dysbiosis in a global and precise manner. Of course, more experiments should be performed on CRC and other intestinal diseases in order to further validate our results.

We are the first to take advantage of metagenomics-data-derived HDCs and used it as a valid indicator of patient stratification via meta-analysis. *In silico* removal of host DNAs from metagenomics data is a recommended procedure [31], however, a quantitative definition of a contaminated sample is still illusive. Our results indicate that the metagenomics data can validate itself by looking at the correlated changes in HDC and related gut microbial species: a sudden increase in HDC without matching alterations in related bacteria is a strong indication of contamination. This line of reasoning can be applied to any host-produced molecules identified from feces, such as DNA, RNAs, proteins, metabolites and even cells, and would pave the way for extracting more host related information directly from fecal samples using multi-omics techniques and making use of them without worrying too much about contamination. As we have shown in this study, host related information directly extracted from fecal samples is reliable and useful.

## Conclusions

In summary, we found that compromised intestinal barrier, as indicated by elevated human DNA contents (HDCs) in feces, is a common feature of intestinal diseases; HDC could be a promising biomarker for intestinal diseases because it signified the abundances changes of most of disease-related species, ranked as the top contributor to machine learning models for patient stratification, and was a better biomarker for effective treatment than fecal calprotectin (FCP). These results suggested that host derived contents may have greater impact on gut microbiota, and called for more attentions to be paid to possible impacts of host derived contents to the gut, the underlying mechanisms, and their possible roles in intestinal diseases, as most of current research focused on the impacts of gut contents to the host.

## Methods

### Metagenomics data analysis

A total of 354 CRC patients, 87 CD patients and 382 controls from eight fecal metagenomics datasets were included in this study. More details, including the nationality and age of these subjects, can be found in Table S1. To remove adapters and low quality of bases, raw reads were filtered and trimmed by Trimmomatic [32] v3.6, using the Truseq3 adapter files and option with MINLEN cutoff 50. To estimate the human DNA contents (HDC) in metagenomics sequencing reads, the remaining reads (clean reads) were aligned to the human reference genome (hg19) using bowtie2 [33] (version 2.3.4.3); the HDC of a sample was calculated as the percentage of mapped reads out of total clean reads in the sample. The human DNA contents measured by quantitative PCR (QPCR) results in CD dataset were obtained from the corresponding publication by Lewis and colleagues [13].

Reads mapped to the human genome were removed before subsequent analyses. Taxonomic abundances of all metagenomic samples were quantified using MetaPhlAn2 [34]. HUMAnN2 [35] was used to calculate relative pathway abundances via mapping reads to ChocoPhlAn database and full UniRef90 database.

In each project, species with max abundance <1% in all samples as well as species whose average abundance across all samples below 0.01% were removed from further analyses. Similarly, pathways with maximum relative abundances less than 1× 10^−6^ in all samples of a project were also removed.

### Statistics and modeling

All processed data were loaded into R (https://www.r-project.org) and analyzed. Wilcoxon Rank Sum Test was used to detect significant between-group differences in relative abundances of taxonomic- and pathway-features; features with P-value < 0.05 were deemed significant. Spearman correlation was used to find HDC correlated species and pathways, features with P-value < 0.05 were selected as significantly correlated features.

The SIAMCAT package (https://bioconductor.org/packages/SIAMCAT/) in R was used to build mathematic classification models (classifiers) that are capable of distinguishing patients and tumor-free participants, extract features that can be used to discriminate different phenotype groups and calculate feature importance scores. Random forest algorithm implemented in SIAMCAT was used for model training and classification.

For the CD data, 20 times repeated ten-fold cross-validation (200 models would be obtained) implemented in the SIAMCAT package was used to assess the within-dataset accuracy of the resulting classifiers. For the CRC data, a so-called leave-one-dataset-out (LODO) analysis was also performed in order to evaluation cross-study performance of the obtained classifiers. In LODO analyses, all datasets except the one used for model testing were pooled as a training dataset which would be implemented the within-dataset ten-fold cross-validation; LODO was performed for each dataset in turn and were repeated 10 times, for all the seven CRC datasets. The LODO training dataset prediction accuracy was measured through ten times repeated ten-fold cross-validation.

## Data Availability

all data are publicly available. the corresponding links to public database have been included in the manuscript.

## Declaration

### List of abbreviations

CIB: Compromised intestinal barrier
HDC: Human DNA content
CRC: Colorectal cancer
CD: Crohn’s disease
IBD: Inflammatory bowel disease
LODO: Leave-one-dataset-out
QPCR: Quantitative polymerase chain reaction
AUC: Area under the receiver-operating characteristics curve

## Ethics approval and consent to participate

From public database and original researches we downloaded raw sequencing data and metadata, which were approved by their corresponding ethics committee. Except total human DNA abundances served as a proxy of shedding cells from human and present in stools, we didn’t use any other information of human genome, such as mutations and gene abundances.

## Consent for publication

Not applicable

## Availability of data and material

Raw sequencing reads and metadata of the seven human colorectal cancer (CRC) metagenomics datasets were obtained from European Nucleotide Archive (ENA) under the following ENA project identifiers: PRJEB10878 [11], PRJEB27928 [15], PRJEB7774 [12], PRJEB12449 [14], and PRJEB6070 [10], PRJNA447983 [16] (cohort 1), and PRJDB4176 [15]. Raw sequencing reads and metadata of the Crohn’s disease metagenomics dataset were obtained from NCBI SRA database under SRA ID: SRP057027 [13].

All processed data and R codes are available at https://github.com/evolgeniusteam/HumanDNAContents_in_CRC_gut_metagenomics; also available are instructions for users to reproduced all our analyses, including figures, supplementary figures and statistics.

## Authors’ contributions

W.H.C. and S.X. designed the study. P.J. collected and analyzed the CRC data, S.L. collected and analyzed the Crohn’s Disease data. S.W. coordinated the data downloads and analysis. X.M.Z contributed to the mathematical modeling and interpretation. W.H.C., P.J. and S.X. wrote the manuscript with all authors contributing to the writing and providing feedbacks. All authors read and approved the final version of the manuscript.

## Acknowledgements

Not applicable

## Additional files

**Table S1**. A list of CRC and CD projects used in this study and the numbers of controls and cases.

**Table S2**. Metadata of participants in CRC projects and their HDC%. HDC means the human DNA content.

**Table S3**. Species that correlated with HDC in more than two CRC datasets statistically. Species whose Spearman p-value <0.05 in any project were deemed as HDC correlated species. The column overlap = “1” means that the specie is a diagnostic feature in other meta-analyses (pmid: 30936548 and 30936547).

**Table S4**. Pathways that correlated with HDCs in more than three CRC datasets. Pathways whose Spearman p-value <0.05 in any project were deemed as HDC correlated pathways. The column overlap =“1” means the pathway is a diagnostic feature in other meta-analysis (pmid: 30936548).

**Table S5**. A list of samples of the CD project and their HDC% produced by two ways. Our HDC% were generated using Bowtie2, while Lewis’s HDC% were generated using Bmtagger (pmid :26468751).

**Table S6**. Species that correlated with HDCs in CD. We calculated Spearman correlation between HDC and species relative abundance in each stage of patients and controls.

**Table S7**. Differential species in the Control+Baseline group of CD.

**Table S8**. Overlapped features of the top 20 important features between CD and CRC. The columns “Importance_in_CRC” and “Importance_in_CD” mean the median importance scores of the top 20 features in the CRC models and CD models, respectively. The columns “ranking_in_CRC” and “ranking_in_CD” mean the importance degrees of the shared features in the CRC models and CD models separately.

**Figure S1**. Log10-transformed relative abundance distribution of all HDC-correlated species after diet and antibodies intervention in CD dataset.

**Figure S2**. Correlation between HDC and fecal calprotectin in CD dataset

**Figure S3**. Correlations between fecal calprotectin and species in CD dataset.

